# Multi-Ancestry GWAS of Neuroticism Identifies Novel Loci and Enhances Fine-Mapping Resolution

**DOI:** 10.64898/2025.12.12.25342160

**Authors:** Mary Kaka, Apostolia Topaloudi, Guangxin Chen, Priya Gupta, Daniel F. Levey, Boran Gao, Peristera Paschou

## Abstract

Neuroticism is a core personality trait linked to emotional instability and increased risk for anxiety, depression, and schizophrenia. Most genome-wide association studies (GWAS) have focused on European populations, limiting the discovery of ancestry-specific genetic influences. To address this gap, we conducted the most diverse multi-ancestry GWAS meta-analysis of neuroticism to date, analyzing 668,780 individuals across African (AFR), European (EUR), East Asian (EAS), and South Asian (SAS) populations using UK Biobank (UKB) and the Million Veterans Program (MVP). We identified seven novel loci through SNP-based GWAS and another three novel genes through gene-based analysis. Multi-ancestry fine-mapping improved causal variant resolution, reducing credible set size by 61%, with 156 putatively causal SNPs, 129 of which were specific to EUR. Tissue enrichment highlighted brain, nerve, and pituitary, pointing to the involvement of GABAergic neurons. Many loci overlap with genes previously associated with psychiatric and cognitive traits, reinforcing neuroticism’s pleiotropic and transdiagnostic nature. Our findings highlight the urgent need to expand genetic diversity in psychiatric genomics, ensuring that precision medicine advances benefit all populations equitably.

## INTRODUCTION

Neuroticism is a fundamental personality trait with profound implications for mental health and overall life quality ^1^. It is characterized by heightened emotional reactivity and a predisposition to negative affect, making it a significant risk factor for various neuropsychiatric disorders, including schizophrenia, anxiety disorders, and depression ^2–5^. As one of the Big Five personality traits, neuroticism plays a crucial role in moderating extraversion and conscientiousness, influencing self-control, and serving as an endophenotype for other brain-related traits^1,6^. Given its strong links to global public health concerns, understanding its genetic architecture is critical for advancing psychiatric genomics and precision medicine.

Twin and family studies estimate that neuroticism is highly heritable (∼40%) ^2–5^, with SNP-based heritability varying from 6-10% ^2–5,7^. Early genome-wide association studies (GWAS) identified several genetic loci associated with neuroticism, including MAPT, NSF, WNT3, CRHR1, and KANSL1 ^2–5^. As sample sizes increased, so did the number of implicated loci, culminating in a recent large-scale GWAS, which analyzed 623,482 individuals from the Million Veterans Program (MVP) and UK Biobank (UKB)^7^. This study identified 216 independent genomic risk loci, replicating previous associations and revealing novel insights into the genetic basis of neuroticism.

Despite these advances, the vast majority of GWAS for neuroticism have been conducted in populations of European descent. Only the most recent efforts have included individuals of African ancestry, and even then, sample sizes remain limited compared to those of European descent ^7^. Critically, no large-scale GWAS has systematically incorporated individuals of East and South Asian ancestry, leaving major gaps in our understanding of the genetic architecture of neuroticism across diverse populations. This lack of representation restricts the generalizability of findings, limits fine-mapping resolution, and hinders the identification of ancestry-specific risk factors^8^. Furthermore, given the greater genetic diversity within African populations, inclusion of these ancestries could significantly enhance genetic discovery and fine mapping efforts by leveraging differences in linkage disequilibrium (LD)^9^. While previous GWAS have largely relied on single-ancestry fine-mapping approaches, most recent multi-ancestry fine-mapping frameworks enhance causal variant resolution, addressing ancestry-specific LD differences and improving functional interpretation^10,11^.

To address these critical gaps, we conducted the most diverse multi-ancestry GWAS meta-analysis of neuroticism to date, analysing 668,780 individuals, and including expanded cohorts of African-ancestry individuals as well as integrating the first large-scale analysis of East and South Asian populations. Through multi-ancestry fine-mapping studies, we identified both ancestry-specific and shared causal risk loci, improving resolution for causal gene discovery. Additionally, extensive post-GWAS analyses provided deeper insights into the biological mechanisms underlying neuroticism. Our results significantly extend previous findings and emphasize the importance of genomic diversity in psychiatric genetics, paving the way for more inclusive and precise models of neuroticism’s genetic architecture.

## METHODS

### Study Samples and Neuroticism Assessment in UK Biobank

The UK Biobank (UKB) is a large-scale, population-based cohort that includes genetic and phenotypic data from more 500,000 participants recruited in the UK^12^. The cohort is predominantly composed of individuals of European descent, with over 90% self-identifying as White British, while the remaining participants report Asian, Black, Mixed, Chinese, or other ancestry. In this study, neuroticism was measured using 12 dichotomous (“yes”/“no”) items from the Eysenck Personality Questionnaire-Revised Short Form (EPQ-RS) ^13^. A detailed description of how the neuroticism score was constructed is provided in the Supplementary Methods, Supplementary Table 1, and its distribution is shown in Supplementary Figure 1.

### Quality Control, Imputation, and Ancestry-Specific GWAS

The initial UKB dataset included 502,490 individuals who underwent quality control following the Ricopilli pipeline^14^, as detailed in the Supplementary Methods. Details about the number of samples analyzed in each population and GWAS are shown in Figure 1 and Supplementary Table 2. Since this study focused on ancestry-specific analyses, participants were categorized into African (AFR), East Asian (EAS), and South Asian (SAS) ancestry groups based on principal component analysis (PCA) using TeraPCA^15^, incorporating the 1000 Genomes (1kG) reference panel (Supplementary Figure 2, Supplementary Table2). Genotype imputation was performed using the TOPMed reference panel via the Michigan Imputation Server^16,17^. Variants with a minor allele frequency (MAF) greater than 1% and an imputation accuracy of r² > 0.7 were retained for downstream analyses, resulting in approximately 11 million SNPs. Genome-wide association analyses (GWAS) were conducted separately within each ancestry group using PLINK 1.9^18^, adjusting for sex, age, and the first three principal components to account for population structure. The European ancestry group was excluded from the individual-level GWAS, as summary statistics for neuroticism were already available from a previous study^2^.

**Figure 1:**
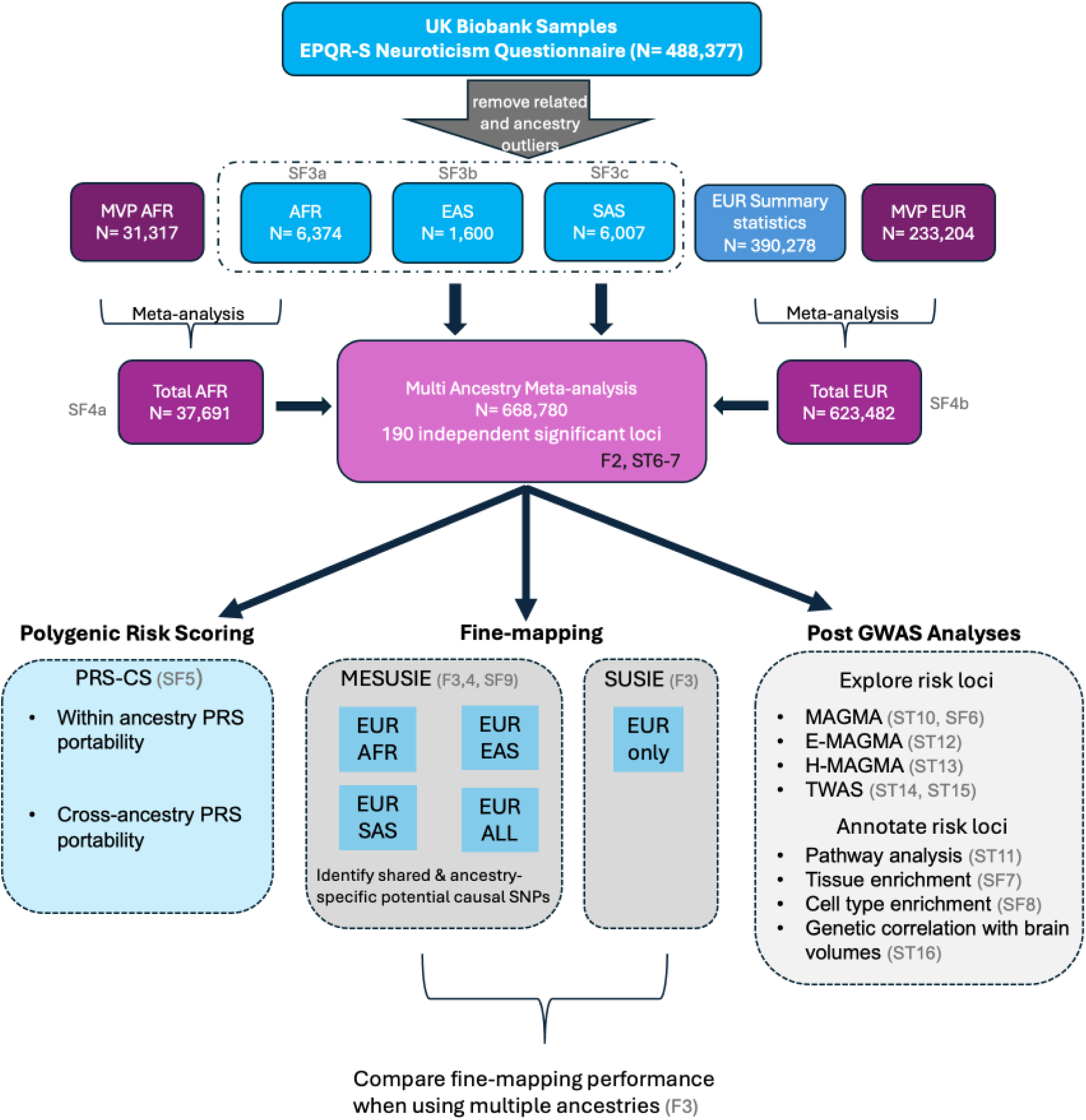
Methodology Flowchart: We first conducted ancestry-specific GWAS for neuroticism in UKB individuals of AFR, SAS, and EAS ancestries. Next, we incorporated AFR and EUR individuals from the MVP sample, followed by a multi-ancestry GWAS meta-analysis across all four populations (AFR, EAS, SAS, EUR). The results of this meta-analysis served as the basis for fine-mapping to identify causal variants, as well as post-GWAS analyses to reveal insights into the functional aspects of neuroticism risk loci. Tables and figures displaying results for each approach are highlighted in light gray.

**Figure 2:**
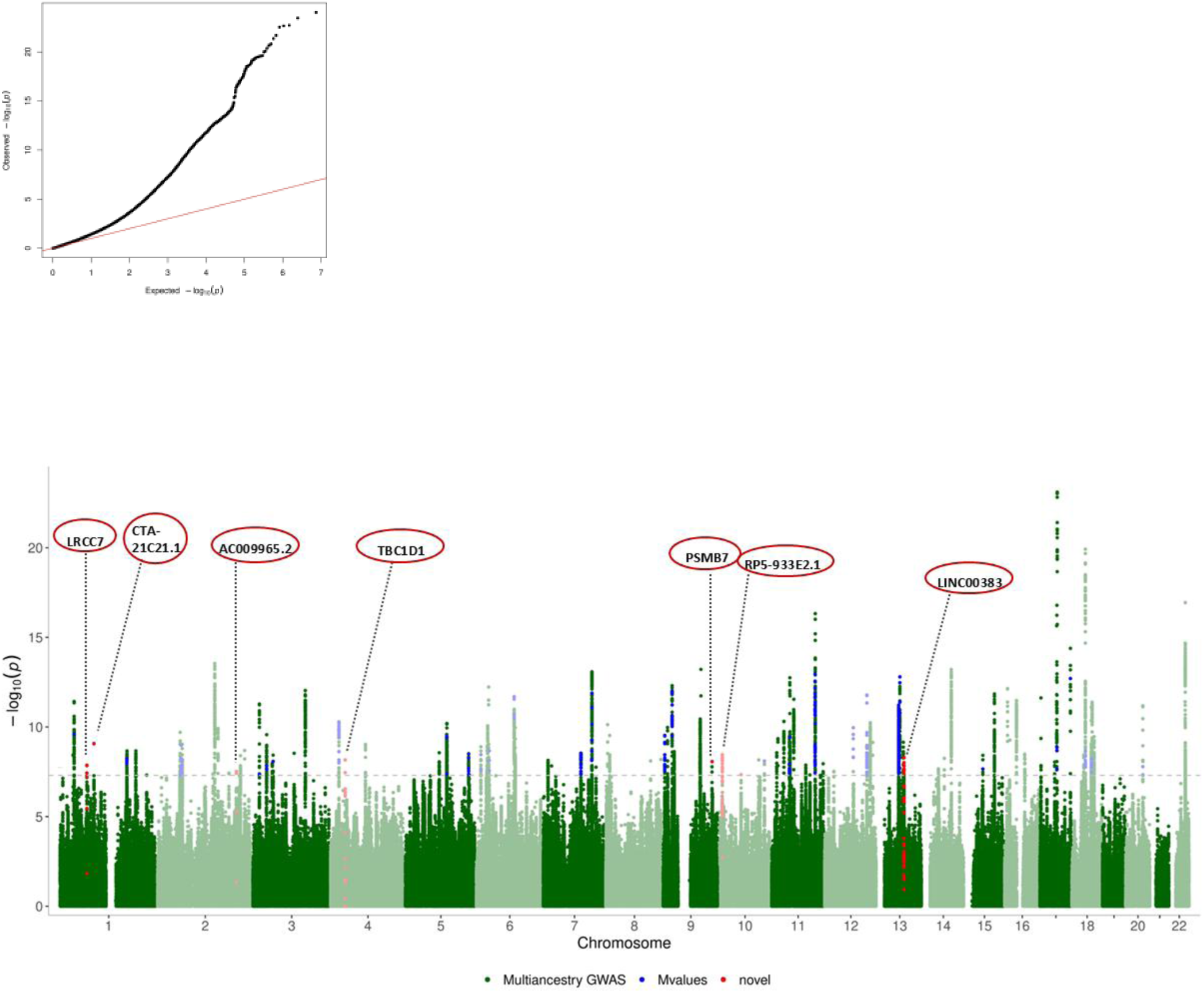
Manhattan and QQ plot of neuroticism multi-ancestry GWAS meta-analysis. 668,780 individuals from African (AFR), European (EUR), East Asian (EAS), and South Asian (SAS) populations were analyzed. Analysis was performed using the Random effects model in METASOFT. The gray dashed line indicates the genome wide significant threshold of p< 5×10^−8^. Shared SNPs between AFR and EUR (mvalues > 0.9) are annotated with blue dots while the tracing gray lines are showing the mapped genes. Red circles indicate seven novel significant genes for neuroticism (Table 1).

**Table 1:** Novel neuroticism loci identified from multi-ancestry GWAS and gene-based analyses. Table (a) shows all 7 novel loci from the multi-ancestry GWAS, while Table (b) shows the 3 novel genes from the MAGMA gene-based analysis.

| CHR | START | END | Lead SNP | P | NEAREST GENE (distance in bp) |
| --- | --- | --- | --- | --- | --- |
| 1 | 70333405 | 70334359 | rs2137591 | 1.38E-08 | LRRC7 (0) |
| 1 | 88828592 | 88828592 | rs2207286 | 8.60E-10 | CTA-21C21.1 (49607) |
| 2 | 204994019 | 204997036 | rs10932046 | 3.08E-08 | AC009965.2 (22464) |
| 4 | 37984238 | 37999855 | rs13111793 | 6.90E-09 | TBC1D1 (0) |
| 9 | 127188174 | 127188174 | rs12683568 | 8.61E-09 | PSMB7 (10451) |
| 10 | 9955322 | 9963119 | rs12572399 | 3.52E-09 | RP5-933E2.1 (139074) |
| 13 | 69802136 | 69878353 | rs12584033 | 5.06E-09 | LINC00383 (9072) |

(b)
| CHR | START | END | P | GENE | Associated traits in GWAS catalog |
| --- | --- | --- | --- | --- | --- |
| 1 | 44679104 | 44686351 | 6.13E-07 | DMAP1 | - |
| 5 | 64859063 | 64883376 | 2.71E-06 | PPWD1 | Brain morphology, Corneal resistance factor, Thalamic Nuclei volume |
| 11 | 58169938 | 58170882 | 2.39E-06 | OR5B3 | Protein quantitative trait loci (liver) |

### Ancestry-Specific and Multi-Ancestry GWAS Meta-Analysis

Full details about the number of samples and SNPs in ancestry-specific and multi-ancestry GWAS as well as comparison to previous GWAS are shown in Supplementary Table 2. To maximize power for detecting signals within each ancestry, a meta-analysis was conducted by combining MVP and UKB data separately for EUR and AFR populations. Although, in Gupta et al.^7^ a meta-analysis of MVP+UKB EUR was performed, we decided to repeat this step excluding heterogeneous SNPs as outlined below. For UKB EUR, summary statistics were obtained from Nagel et al.^2^, while for UKB AFR, the results from the ancestry-specific GWAS were used. A fixed-effects meta-analysis was performed using METAL^19^, with SNPs exhibiting a heterogeneity p-value below 0.05 excluded. For MVP EUR and AFR, we obtained summary statistics from Gupta et al.^7^. After meta-analysis, the final sample sizes included 37,691 individuals of African ancestry and 623,482 individuals of European ancestry. A detailed description about the MVP cohort, as well as information on the genotyping, imputation, quality control, and GWAS is described in Gupta et al ^7^.

In order to detect signals across diverse ancestries and account for heterogeneous effect sizes, we further conducted a multi-ancestry meta-analysis focusing on SNPs that were common across AFR, SAS, EAS, and EUR using METASOFT’s random-effects model [Han and Eskin’s (RE2) model]^20^, which is more effective in detecting signals under effect size heterogeneity. See Supplementary Table 2 for details about the number of samples and SNPs in the analysis. In addition, we estimated the posterior probability (m-value) to identify SNPs with heterogeneous effect sizes across ancestries (m-value>0.9) . Then we defined LD independent risk loci and compared with previous literature to identify novel risk loci. The details of how the independent risk loci and novel loci are identified are described in Supplementary Methods. The methodology workflow is shown in Figure 1.

### Fine-Mapping Analysis

Fine-mapping analyses were conducted to refine putative causal variants associated with neuroticism. Single-ancestry fine-mapping was performed using SuSiE^21^ in the European ancestry group, which had the largest sample size, while multi-ancestry fine-mapping was conducted using MESuSiE^10^ to leverage genetic diversity across populations. Fine-mapping analyses were conducted separately for single-ancestry (EUR) and multi-ancestry models, including all possible ancestry pair combinations. The independent risk loci defined from the multi-ancestry GWAS meta-analysis were used as input for fine-mapping (See **Supplementary Methods**). LD reference panels used in fine-mapping were derived from in-sample UKB genotype data for each ancestry group. A putative causal SNP was defined as one with a posterior inclusion probability (PIP) greater than 0.5. The resolution of fine-mapping was further assessed by comparing the sizes of 99% credible sets.

### Within- and Cross-Ancestry Polygenic Risk Score (PRS) Analysis

The portability of neuroticism polygenic risk scores (PRS) across ancestries was evaluated using PRS-CS^22^, applying the UKB LD reference panel. PRS was calculated using PLINK and assessed in both within- and cross-ancestry settings. For within-ancestry PRS, MVP and UKB EUR samples were used as the discovery and target datasets, respectively, while MVP and UKB AFR samples were used for within-African ancestry validation. Cross-ancestry PRS transferability was tested by using EUR (MVP + UKB) summary statistics as the discovery dataset and AFR, SAS, and EAS UKB samples as the target populations. PRS distributions were divided into quintiles, and associations with neuroticism were tested using linear regression models, adjusting for sex and ancestry-related covariates using R.

### Post-GWAS Functional and Enrichment Analyses

To functionally characterize GWAS loci and identify biologically relevant pathways, four complementary approaches were applied: genome-wide gene-based association analysis using MAGMA^23^, expression quantitative trait locus (eQTL) mapping using e-MAGMA ^24^, chromatin interaction mapping using h-MAGMA^25^, and transcriptome-wide association studies (TWAS)^26^. Multiple testing correction was applied using the Bonferroni method. Loci were classified as novel if they had not been previously reported in neuroticism GWAS or in the GWAS Catalog.

To assess tissue-specific associations, gene expression enrichment analysis was conducted using MAGMA in FUMA^27^. This included 30 broad and 54 specific tissue types, with significance determined using Bonferroni correction. Cell-type-specific enrichment was analyzed using FUMA’s cell-type module, selecting brain-related datasets (Supplementary Table 3). Gene ontology (GO) analysis was performed using g:Profiler^28^ to identify enrichment of biological processes, cellular components, and molecular functions among associated genes.

### Genetic Correlation with Brain Volume Traits

Genetic correlations between neuroticism and seven subcortical brain volume traits from the ENIGMA consortium ^29^ were estimated using LD Score regression^30^. Given the high heritability of brain volume traits^31^ and their potential genetic overlap with personality traits^32^, this analysis aimed to explore shared genetic architecture between neuroticism and structural brain variations. Since both neuroticism and brain volume traits were analyzed in multi-ancestry cohorts, the corresponding LD scores are derived from the LD reference panel constructed with multi-ancestry participants reflecting population proportions in the neuroticism GWAS (See **Supplementary Methods**). Bonferroni correction was used for multiple testing adjustments.

## RESULTS

### GWAS on UKB Individuals of African, East Asian, and South Asian Ancestries

We conducted ancestry-specific GWAS for neuroticism in UKB participants, analyzing 6,374 individuals of African ancestry (AFR), 6,007 South Asians (SAS), and 1,600 East Asians (EAS). No variants reached genome-wide significance (p<5×10⁻⁸) in UKB AFR, SAS, or EAS GWAS (Supplementary Figure 3a-c). To increase statistical power for AFR samples, we integrated AFR summary statistics from the MVP cohort, yielding a total sample of 37,691 individuals of African ancestry. Despite the significantly expanded dataset compared to Gupta et al.^7^, still no SNPs achieved genome-wide significance (Supplementary Figure 4a). However, at a suggestive threshold (p < 10⁻⁵), we identified 52 variants in AFR, 24 in SAS, and 22 in EAS associated with neuroticism (Supplementary Table 4). A list of top-scoring independent SNPs from AFR (MVP+UKB), EAS, and SAS GWAS is provided in Supplementary Table 3. In contrast, the meta-analysis of EUR UKB and MVP data (623,482 individuals) identified 245 genome-wide significant loci (Supplementary Figure 4b). As detailed in Methods, we applied a stricter QC approach, removing SNPs with high heterogeneity and implementing more stringent QC measures. This led to the identification of nine fewer significant loci compared to the Gupta et al. study^7^. See Supplementary Table 5 for full comparison.

**Figure 3:**
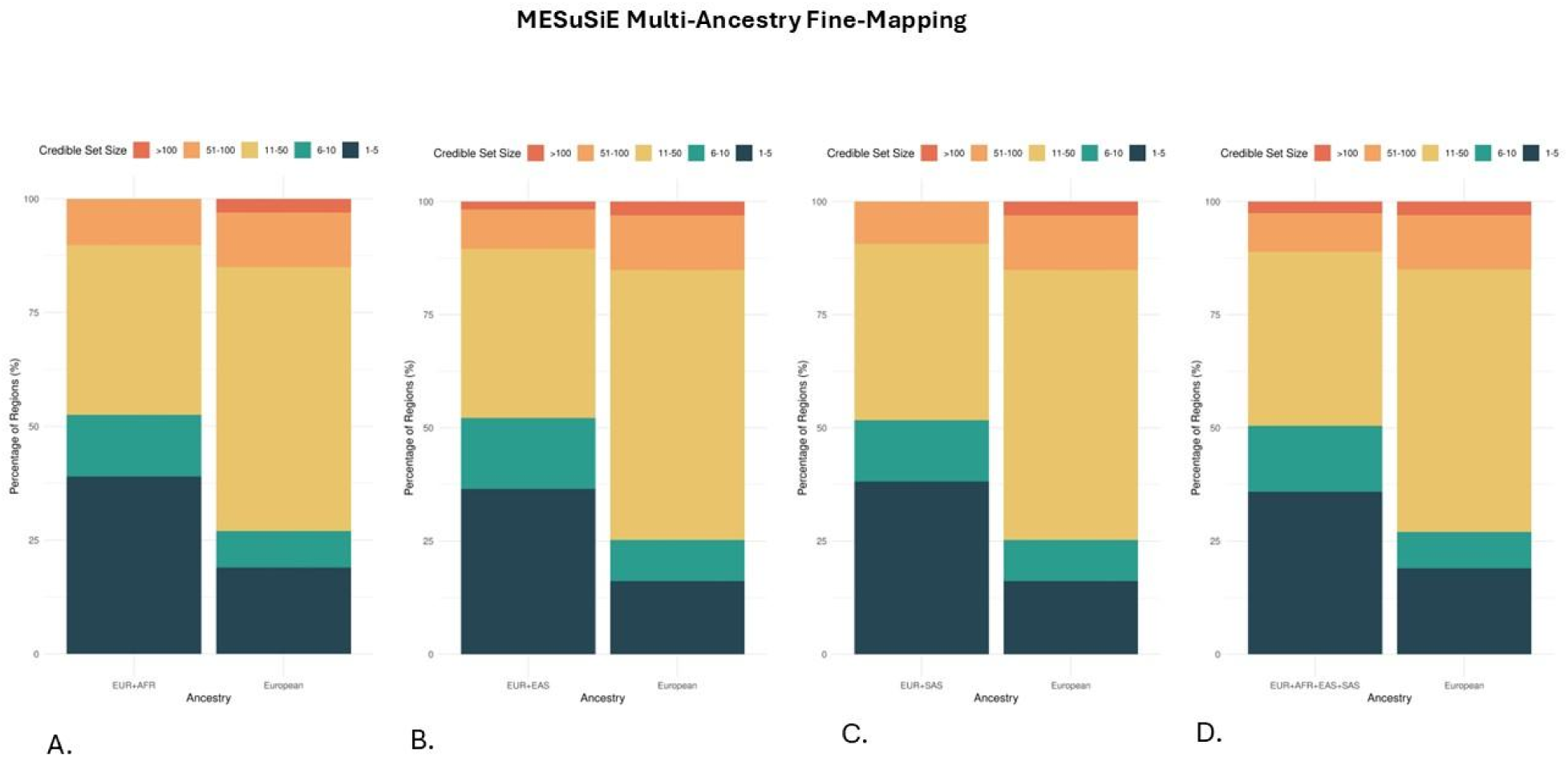
Reduction in Credible Set Sizes Using Multi-Ancestry Fine-Mapping Compared to European-Only Analysis. Bar charts comparing credible set sizes in European-only (SuSiE) versus multi-ancestry fine-mapping (MESuSiE) across 136 genomic regions. Each pair of bars represents the number of credible set variants identified in the European-only fine-mapping (left) versus the corresponding multi-ancestry fine-mapping approach (right). Multi-ancestry fine-mapping was performed using different population combinations: (a) EUR+AFR, (b) EUR+EAS, (c) EUR+SAS, and (d) EUR+ALL (integrating all ancestries). The results demonstrate the increased resolution of fine-mapping when incorporating genetic diversity, with a reduction in credible set sizes across multi-ancestry analyses.

**Figure 4:**
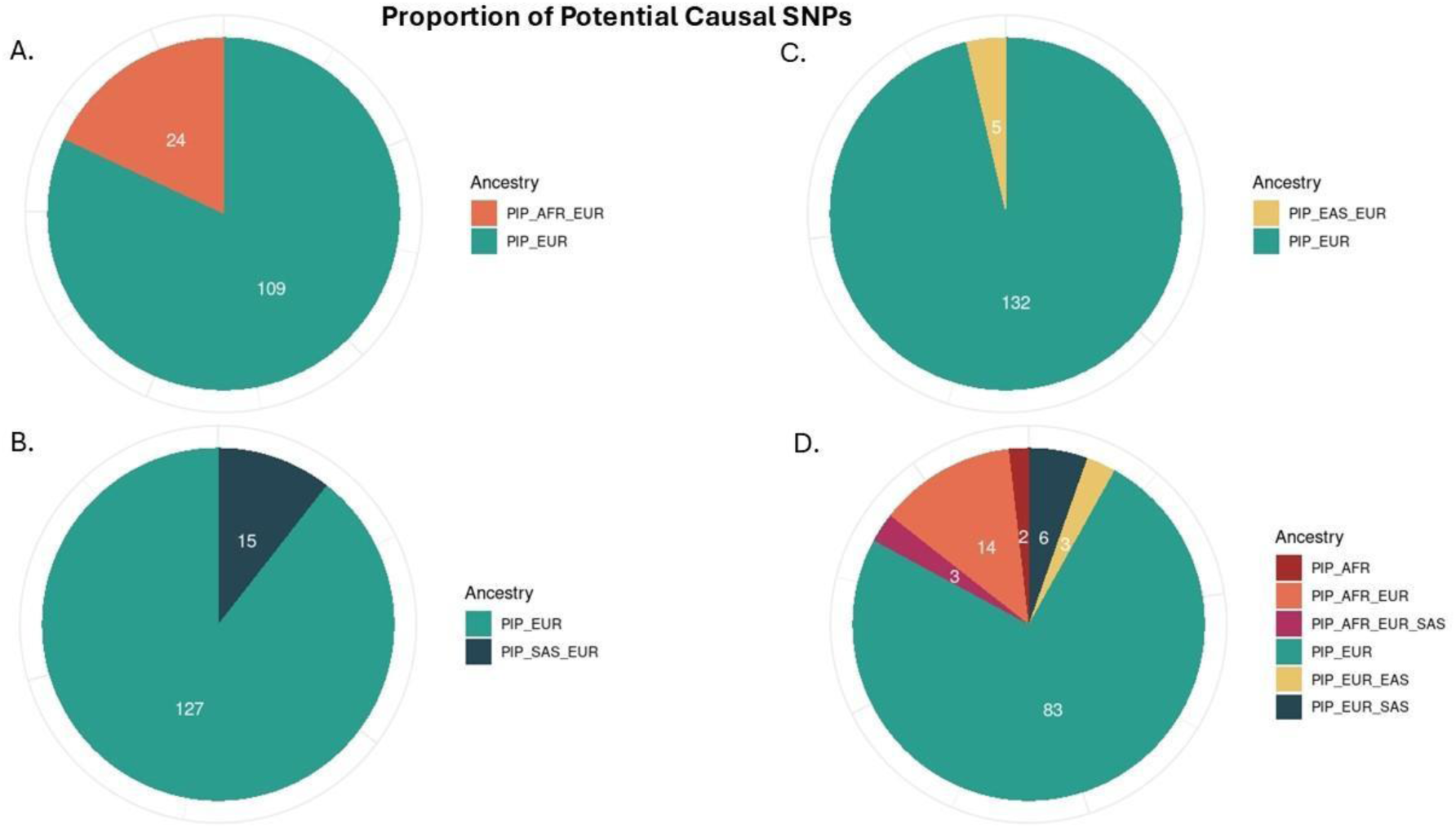
Proportions of Potentially Causal SNPs Identified in Multi-Ancestry Fine-Mapping Compared to European-Only Analysis. Pie charts depicting the distribution of potentially causal SNPs identified through multi-ancestry fine-mapping (MESuSiE) compared to European-only fine-mapping (SuSiE). Each chart represents a different population combination used in multi-ancestry fine-mapping: (a) EUR+AFR, (b) EUR+EAS, (c) EUR+SAS, and (d) EUR+ALL (all populations combined). The segments illustrate the proportion of SNPs retained as potentially causal within each fine-mapping approach, demonstrating how incorporating diverse populations influences SNP prioritization and improves causal variant resolution.

### Multi-ancestry GWAS Meta-analysis for Neuroticism

We integrated the AFR, EUR, SAS, and EAS ancestry-specific GWAS results from UKB and MVP to run the most diverse multi-ancestry GWAS to date and investigate ancestry-shared versus ancestry-specific genetic associations. This analysis included 668,780 individuals, incorporating an additional 13,891 samples as compared to the Gupta et al.^7^, and retained 3,692,041 SNPs after quality control and focusing on SNPs that are common across all datasets (Supplementary Table 2). Association summary statistics from all datasets were meta-analyzed using the random effects model (RE2) in METASOFT, which accounts for effect size heterogeneity across populations. A total of 2,727 genome-wide significant SNPs were identified, with rs199454 emerging as the top variant (p = 7.89 × 10⁻²⁴; BETA = −0.007) (Figure 2). To refine independent association signals, we performed clumping and regional comparison with the previously reported MVP GWAS^7^. This identified 190 independent genome-wide significant loci (Supplementary Table 6), of which seven were novel (Table 1).

To further evaluate ancestry-specific contributions, we estimated posterior probabilities (m-values) for each SNP across populations, identifying loci with m-values > 0.9, indicative of strong ancestry-specific effects. All genome-wide significant SNPs in EUR had m-values > 0.9, reflecting the large sample size of the European dataset. Among these, 595 SNPs also had m-values > 0.9 in AFR, suggesting potential shared genetic effects between African and European populations. However, no SNPs reached this threshold in SAS or EAS, highlighting the reduced power in these populations, likely due to smaller sample sizes (Supplementary Table 7). To further explore population-specific effects, we compared lead SNPs (i.e. top SNPs in each clumped region) from the multi-ancestry meta-analysis with ancestry-specific GWAS results (Supplementary Table 8).

### Multi-ancestry GWAS Fine-mapping for Neuroticism

Next, we pursued fine-mapping with a goal to detect potentially causal SNPs in our multi-ancestry GWAS meta-analysis and investigate ancestry-specific variants. We used a multi-ancestry fine-mapping method (MESuSiE)^10^ to derive 99% credible sets for 136 loci (after excluding genome - wide significant loci that included a single SNP). In order to discern cross-ancestry effects, we performed pairwise fine-mapping of EUR and each of the AFR, SAS, and EAS separately, as well as all four populations combined. We also implemented single-ancestry fine-mapping of the same loci based on GWAS conducted in participants of European ancestry, using SuSiE, to compare the performance of fine-mapping on European and multi-ancestry samples.

In all cases, integrating non-european populations led to improved fine-mapping resolution. Among the 136 loci we fine-mapped, 83 loci (61%) showed a smaller 99% credible set in the multi-ancestry fine-mapping (FMAP_ALL) compared to the European-only (EUR) fine-mapping, indicating higher resolution in the multi-ancestry analysis. When the AFR populations were integrated in analysis, the median size of the 99% credible sets was reduced from 13 to eight variants in the multi-ancestry versus EUR-only fine-mapping (FMAP_AFR) (Figure 3a). It was similarly reduced from 14 to eight variants using EAS (FMAP_EAS) and SAS (FMAP_SAS) (Figure 3b,3c), and dropped from 13 to eight variants using all populations (FMAP_ALL)(Figure 3d) in the multi-ancestry fine-mapping. This drop reflects the increased resolution enabled by differences in linkage disequilibrium across ancestries. A total of 156 SNPs across all four sample combinations we interrogated in our multi-ancestry fine-mapping analyses (FMAP_AFR, FMAP_EAS, FMAP_SAS, FMAP_ALL) were found to be potentially causal (PIP>0.5). We also found that 129 (82.7%) of SNPs identified as potentially causal were specific to European ancestry (Supplementary Table 9; Figures 4a-d). This apparent enrichment of EUR-specific signals could be largely attributed to the greater statistical power and larger sample size of the EUR cohort, which contributes strongly to overall signal detection.

### Neuroticism PRS Estimations within and across Ancestries

We proceeded to test the predictive accuracy of our neuroticism multi-ancestry GWAS PRS both within and across ancestries. In the case of tests within EUR samples, we observed increasing OR estimates in the fifth versus the first PRS quintile (OR=2.65, 95% C.I. = 2.54, 2.75) (Supplementary Figure 5). On the contrary, we did not observe the same pattern in AFR, EAS, and SAS samples. This could be attributed to lower sample sizes and GWAS power in these populations as compared to EUR but also points to the high genetic diversity of AFR populations. We also observed low ORs in all quantiles when attempting to perform cross-ancestry tests, indicating low PRS portability.

### Gene-Based Associations, Tissue-Specific Enrichment, and Functional Insights into Neuroticism

To further interpret the findings from our multi-ancestry GWAS, we conducted gene-based association analysis using MAGMA, which evaluates the joint effects of multiple variants within each gene. Using a multi-ancestry LD reference from UKB, we tested 16,911 genes and identified 119 genes as significantly associated with neuroticism after Bonferroni correction (p < 2.96 × 10⁻⁶). Among the significant genes, three were previously unreported in neuroticism GWAS, including DMAP1 (p = 6.13 × 10⁻⁷), PPWD1 (p = 2.71 × 10⁻⁶), and OR5B3 (p = 2.39 × 10⁻⁶) (Table 1, Supplementary Figure 6, Supplementary Table 10).

We then performed tissue and cell-type enrichment analyses using MAGMA in FUMA to identify biological pathways relevant to neuroticism. Tissue enrichment analysis was conducted on 30 broad and 53 specific tissue types from GTEx. After Bonferroni correction (p < 1.7 × 10⁻³), the Brain, Nerve, and Pituitary tissues showed significant enrichment (Supplementary Figure 7a). Further analysis of specific brain regions revealed 13 significantly associated brain regions (p < 9.4 × 10⁻⁴) (Supplementary Figure 7b). Gene set enrichment analysis using g:Profiler identified 11 significant Gene Ontology (GO) terms linked to neuroticism-associated genes (Supplementary Table 11). To investigate enrichment at the cell-type level, we performed FUMA cell-type analysis using brain-specific datasets (Supplementary Table 3). Among the tested brain cell types, GABAergic neurons from the Linnarsson Human Midbrain dataset showed a significant association with neuroticism. This association remained robust after stepwise conditional analysis and multiple testing correction (p = 1.40 × 10⁻¹¹) (Supplementary Figure 8).

To prioritize genes with functional relevance, we conducted e-MAGMA and h-MAGMA analyses, integrating gene expression (e-MAGMA) and chromatin interaction data (h-MAGMA). e-MAGMA analysis, using eQTL data from 13 GTEx brain tissues, identified 165 genes whose expression levels were significantly associated with neuroticism (Supplementary Table 12), 20 of which were novel. h-MAGMA, which incorporates Hi-C chromatin interaction data, further prioritized 217 genes based on their regulatory roles in brain tissue (Supplementary Table 13), including 81 novel genes. Among the genes prioritized by both methods, ZC3H7B, a known neuroticism associated locus^2,7^, exhibited the strongest association (e-MAGMA: p = 1.15 × 10⁻¹⁷, Brain Frontal Cortex; h-MAGMA: p = 4.72 × 10⁻¹⁵).

To further explore gene expression associations, we conducted Transcriptome-Wide Association Studies (TWAS) using GTEx brain tissue data and the multi-ancestry GWAS summary statistics. After Bonferroni correction for 18,684 tested genes, we identified 111 significant genes, of which 27 were novel (Supplementary Tables 14 and 15). The most significant association was observed for SEMA3B-AS1 (p = 1.96 × 10⁻⁷³), a previously reported neuroticism associated locus^7^.

Lastly, we investigated potential genetic correlations between neuroticism and brain structure, focusing on seven subcortical brain volumes from the ENIGMA consortium^29^. After Bonferroni correction for multiple comparisons, we identified a significant genetic correlation between neuroticism and caudate volume (p = 0.002, rg = 0.19) (Supplementary Table 16). This finding suggests potential neuroanatomical links and shared genetic basis between neuroticism and subcortical brain regions involved in emotional regulation and cognitive processing.

### Converging Evidence from GWAS and post-GWAS Approaches

Overall, our multi-ancestry GWAS and post-GWAS revealed more than 500 unique significant genes, thus we analyzed results to identify genes that are supported via multiple approaches. In the GWAS, MAGMA gene-based analysis, e-MAGMA, h-MAGMA, TWAS, and fine-mapping analyses, genes were considered significant using a Bonferroni multiple testing correction approach. Eight genes were found to be significant across all six GWAS and post-GWAS methods (Supplementary Table 17). Out of all 532 implicated genes, we also found 152 genes to be novel not having previously been implicated in neuroticism via GWAS (Supplementary Table 18-20). Taken together, these results point to potential functional links for the GWAS-associated variants and give higher credibility to genes with convergent evidence of association from multiple methods.

## Discussion

In the most diverse multi-ancestry GWAS meta-analysis of neuroticism to date, we analyzed genomic data from over 660,000 individuals across European (EUR), African (AFR), East Asian (EAS), and South Asian (SAS) populations, expanding the previous GWAS by 14,000 non-european samples and identifying multiple novel genome-wide significant loci. We note that this is the first study to incorporate individuals of Asian ancestry in a neuroticism GWAS, and fine-mapping analyses. We uncovered both shared and ancestry-specific loci with potential causal relevance, emphasizing the power of genetic diversity in refining risk loci. Notably, the reduction in credible set sizes in multi-ancestry fine-mapping underscores the impact of genetic diversity in pinpointing causal variants^11^. Our findings highlight the importance of expanding diverse genomic datasets to improve the accuracy, generalizability, and translational impact of genetic discoveries in neuropsychiatric research.

Out of 190 independent genomic loci that we found to be significantly associated with neuroticism in our multi-ancestry GWAS, seven were novel (LRCC7, CTA-21C21.1, AC009965.2, TBC1D1, PSMB7, RP5-933E2.1, LINC00383). Among these novel loci, several genes have been previously implicated in brain-related pathways and traits. LRRC7 is a postsynaptic scaffolding protein essential for synaptic plasticity and neurotransmission, particularly through its interactions with NMDA receptors and CaMKIIα^33^. Variants in LRRC7 have been associated with emotional dysregulation, ADHD, autism spectrum disorder (ASD), and aggression, suggesting a role in stress susceptibility and emotional reactivity^34^. As for CTA-21C21.1 (Cancer-Testis Antigens), it is a tumor gene present in the germ cells of testes, ovaries and trophoblasts, and has been found to undergo deregulated expression in tumor and malignant cells. CTA genes are either X-linked or autosomal, favorably expressed in spermatogonia and spermatocytes, respectively^35^. PSMB7, a core component of the proteasome complex^36^, is crucial for protein degradation and neuronal homeostasis, and while its direct link to neuroticism remains unclear, proteasome function is critical for stress resilience and synaptic maintenance^37^. TBC1D1 has been implicated in obesity and related traits ^38^, and has been reported to be significantly associated with smoking initiation in the GWAS Catalog ^39^. As for LINC00383, AC009965.2 and RP5-933E2.1, they are long interspersed non-coding (LINC) RNA genes. Hence, our findings provide new candidate genes for future functional studies.

The gene-based analysis aggregating the SNP effect into genes identified three novel loci DMAP1, PPWD1, and OR5B3 which warrant further investigation. DMAP1 assists DNMT1 to facilitate DNA methylation ^40^, and is also involved in histone acetylation, both key processes in epigenetic gene regulation ^41^. Although there is evidence DNMTs including DNMT1 are actively involved in brain gene expression ^42^, there are no studies yet supporting a direct DMAP1 involvement. PPWD1 is involved in protein folding, acting as PPIase (peptidyl-prolyl cis–trans isomerase) that serve as catalysts in oligopeptides ^43^. It has also been assumed to be involved in mRNA splicing ^44^. Furthermore, PPWD1 has also been reported in the GWAS Catalog to be associated with brain morphology, corneal resistance factor, and thalamic nuclei volume ^45,46^; ^47^. Finally, OR5B3 encodes an olfactory receptor which is responsible for odor detection by binding odorant molecules in the nose and initiating neuronal responses ^48^.

In order to further investigate the biological and functional underpinnings of neuroticism, we integrated the results from the gene-based analysis with downstream analyses. Accounting for multiple layers of information, such as gene expression and hi-C data, we sought to unravel the functional relevance of associated loci ^49^. Our tissue and cell-type enrichment analyses revealed significant associations with the brain, pituitary, and GABAergic neurons. Disruptions in GABAergic signaling have been implicated in several anxiety and mood disorders, such as generalized anxiety disorder and depression, which are core elements of neuroticism ^50,51^.

We found *eight genes* to be significant across all six of our GWAS and post-GWAS analyses, making them strong candidates for further investigation. These genes are *AREL1, ZC3H7B, MED19, SLC39A13, KCTD10, RANGAP1, EP300, DLST* and have been previously reported in neuroticism GWASs ^2,52,53^. Their significance across a variety of methods that we implemented in our analysis, including transcriptomic and functional approaches, highlights their robustness and potential involvement in biological processes that underlie neuroticism.

Additionally, across all GWAS and post-GWAS investigations, we found a total of 152 genes (in at least one analysis) that had not been previously implicated in neuroticism GWAS and warrant further investigation. Several of these genes have been previously associated with psychiatric disorders and behavioral traits, reinforcing the genetic overlap between neuroticism and neuropsychiatric conditions ^54^. For instance, TCF4-AS1, POU3F2 , MIR9-2, and FURIN ^55–57^ have been associated with schizophrenia, bipolar disorder, and major depressive disorder, highlighting shared neurodevelopmental pathways. Genes such as TBC1D1, FLOT1, and PDE1C have been associated with smoking initiation, risk-taking, and addiction^39,58,59^, while MCHR2 and USP4^39,56^ connect neuroticism to ADHD and mood instability. Additionally, HLA-DRB6 and PSTPIP1^60,61^ suggest a role for immune-brain interactions in emotional regulation, and genes like TBX^62^ link neuroticism to cognitive function and neurodegeneration. These findings underscore the pleiotropic nature of neuroticism-associated loci, supporting its role as a transdiagnostic risk factor across personality traits, psychiatric disorders, and cognitive function.

We also observed a significant genetic correlation between neuroticism and caudate volume, highlighting a potential shared genetic basis between this personality trait and subcortical brain structure. Previous studies have established that Schizophrenia, in particular, neuroleptic-naive subjects experiencing their first episode of the disorder, show a reduced volume in the caudate nucleus ^63,64^. Also, it has been found that there are reductions in caudate volume in individuals with Major Depressive Disorders (MDD) as compared with healthy subjects ^65^. More recently, another study showed the association between Neuroticism and the caudate region ^66^. In examining the relationship between gain/loss incentives and changes in brain caudate activation, they found that higher PRS for Neuroticism incurred lesser activation in this brain region.

Although we incorporated the largest available non-EUR datasets in this study, still larger sample sizes would be needed to provide enough power to detect significant loci in the ancestry-specific GWAS. Additionally, many of the datasets for downstream analysis incorporating transcriptomic and hi-C data were mostly from European individuals which potentially limits our ability to accurately assess gene expression and regulation in diverse populations. This work highlights the need for the community to continue to work towards inclusive resources, collection of samples, tools, and reference maps for analysis of diverse populations from around the world.

Our findings highlight the urgent need to expand genetic research beyond European populations to enhance the accuracy, generalizability, and translational impact of psychiatric genomics. As we also show here, current PRS for neuropsychiatric traits, including neuroticism, perform poorly in non-European populations due to population-specific genetic architecture and differences in LD^67^. The negative result we observed in the PRS analysis highlights a key limitation in the current application of PRS and reinforces the need for more inclusive research. Future studies should incorporate larger multi-ancestry training datasets as they become available, as well as improved statistical frameworks to enhance PRS portability across diverse populations. By integrating ancestrally diverse datasets, our study demonstrates how multi-ancestry fine-mapping improves causal variant resolution. Additionally, identifying both shared and ancestry-specific genetic influences enhances our understanding of neuroticism’s biological basis across populations, supporting the development of more inclusive and tailored interventions. Given the transdiagnostic nature of neuroticism-associated loci, these insights are particularly relevant for addressing mental health disparities and ensuring that future advances in precision psychiatry benefit all populations equitably.

## Funding acknowledgements

DFL was supported by VA ORD grant 5IK2BX005058, PP was supported by NSF grants 1715202 and 2006929.

## Data Availability

Information on availability of GWAS summary statistics are listed in their corresponding publications.

