## Supplementary Materials for "Multi-Ancestry GWAS of Neuroticism Identifies Novel Loci and Enhances Fine-Mapping Resolution"

### Table of contents


**Supplementary Methods** **2**

Samples and Neuroticism Definition on UK Biobank 2

UKB Quality Control, Imputation, and ancestry-specific GWAS 2

LD independent genome-wide significant loci and novel loci 3

MAGMA Gene-Based analysis 3

e-MAGMA and h-MAGMA 3

Transcriptome Wide Association Study (TWAS) 4

**Supplementary Figures** **5**

Supplementary Figure 1: Barplot of the Distribution of Neuroticism Scores in UKB 5

Supplementary Figure 2: PCA plot of UKB and 1000 Genomes samples 6

Supplementary Figure 3: Manhattan Plots of Individual GWAS in UK Biobank individuals. 7

Supplementary Figure 4: Manhattan and qq plots of Multi-ancestry analyses 8

Supplementary Figure 5: Within-ancestry and cross-ancestry PRS associations of Neuroticism. 9

Supplementary Figure 6: Manhattan and qq plots of gene-based analysis using MAGMA 10

Supplementary Figure 7: Tissue enrichment analysis in FUMA. 11

Supplementary Figure 8: FUMA Cell-type results. 12

**References** **14**

#### Supplementary Methods

**Samples and Neuroticism Definition on UK Biobank**

The UK Biobank (UKB) is a large-scale, population-based, prospective cohort that recruited over 500,000 participants from the UK aged 40–69 years old. The biobank contains genetic data of the participants and a wide range of phenotypic information [1]. More than 90% of the participants reported White British background, while the rest self-identified as Asian, Black, Mixed, Chinese, or other ethnic backgrounds. The data we used in this study were obtained from UK Biobank under application number #61553.

Neuroticism was measured with 12 dichotomous ("yes"/"no") items of the Eysenck Personality Questionnaire Revised Short Form (EPQ-RS) (Supplementary Table 1), which has been shown to have reliability >0.81[2]. Participants with valid responses to <10 items were excluded from our analyses. A weighted Neuroticism sum-score was then calculated by adding up individual valid item responses, and dividing that sum by the total number of valid responses, as also described in previous study[3]. Higher sum-scores indicate higher levels of Neuroticism (Supplementary Figure 1).

**UKB Quality Control, Imputation, and ancestry-specific GWAS**

The initial UK Biobank dataset included 502,491 individuals genotyped on the Affymetrix UK BiLEVE Axiom array or the Affymetrix UK Biobank Axiom array. Initially, we performed a first quality control (QC) round following the Ricopilli pipeline [4]. Briefly, we excluded individuals with call rate < 0.98, absolute value of inbreeding coefficient < 0.2, genomic sex discrepancy with reported sex, and variants with call rate < 0.98, and Hardy–Weinberg equilibrium P-value < $10^{-6}$. To remove related individuals, we used the provided UKB kinship coefficient <0.0625 (third-degree relatedness [5]). Since we were interested in performing ancestry-specific analyses of Neuroticism, for Africans (AFR), East Asians (EAS), and South Asians (SAS), we performed Principal component analysis (PCA) using TeraPCA [6] including 1000 Genomes (1kG) as a reference to assign the participants in one of the four ethnic groups using an additional layer of information other than the self-reported ancestry. We assigned UKB individuals in an ancestry group if they were within 3 SD from the mean of the 1kG superpopulation for the first 3 PCs, while ensuring consistency with their self-reported ethnicity.

To prepare the ancestry specific datasets for the association tests, we extracted the individuals identified in African, South Asian, and East Asian ethnic groups from the original UKB data, and proceeded to an independent pre-imputation QC with the same parameters as mentioned above. Then, we performed genotype imputation for each dataset using TOPMed [7] as reference in TOPMed imputation server [8], and SNPs with minor allele frequency (MAF) >1% and imputation accuracy of r2>0.7 were included in the downstream analyses. Since there is already available a UKB European ancestry Neuroticism GWAS [9] we did not repeat the analysis for this population.

**LD independent genome-wide significant loci and novel loci**

After obtaining the multi-ancestry summary statistics we performed clumping to define LD independent significant loci, using as index the genome wide significant SNPs (p<5e-8), and including in the region SNPs with r2>0.3 up to 3000 kb away. An LD reference panel reflecting population proportions in the neuroticism GWAS was constructed. A total of 50,000 UKB individuals were randomly selected based on their ancestry distributions: 48,053 of European ancestry, 887 of African ancestry, 223 of East Asian ancestry, and 836 of South Asian ancestry.

For the MVP+UKB summary statistics of EUR, the same clumping parameters are used, with a LD reference panel constructed from randomly selected 50,000 Europeans samples of UKB. These regions were then checked for physical overlap with the largest published Neuroticism GWAS [10], and those not present in the previous study were considered novel.

**MAGMA Gene-Based analysis**

Then, we performed a gene-based association analysis using MAGMA[11] to detect candidate genes associated with Neuroticism. We performed the analyses using the default parameters (annotating SNPs to genes if they physically overlap), and we used a multi-ancestry LD reference from UKB. We applied Bonferroni correction to select significant genes (*p-value* < $3x10^{-6}$ *for 16911 genes*).

**e-MAGMA and h-MAGMA**

To further prioritize genes associated with Neuroticism, we performed e-MAGMA and h-MAGMA, utilizing tissue-specific eQTL information and Hi-C chromatin measurements to characterize risk variants based on their putative genes[12][13]. eMAGMA, modifies the annotation stage of the MAGMA pipeline by mapping SNPs to genes based on tissue-specific eQTL information in GTEx[14], in this study we used 13 brain tissues. For h-MAGMA we used the Hi-C mapping from the adult brain tissues which was provided online by the tool authors[15].

**Transcriptome Wide Association Study (TWAS)**

Next, we performed a Transcriptome Wide Association Study (TWAS) to identify genes whose transcript expression is associated with Neuroticism. For our analysis we utilized S-PrediXcan [16] to perform the tissue specific TWAS of Neuroticism GWAS and eQTL data from 13 GTEx v8 brain tissues. For the analyses we used pre-trained joint-tissue imputation (JTI) [17] prediction models, which improve the prediction performance in the target tissue by borrowing information from different tissues, considering shared genetic effects of regulation between them. Finally, we applied the aggregated Cauchy association test (ACAT)[18] method to calculate the combined p-value of the 13 brain tissues.

### Supplementary Figures

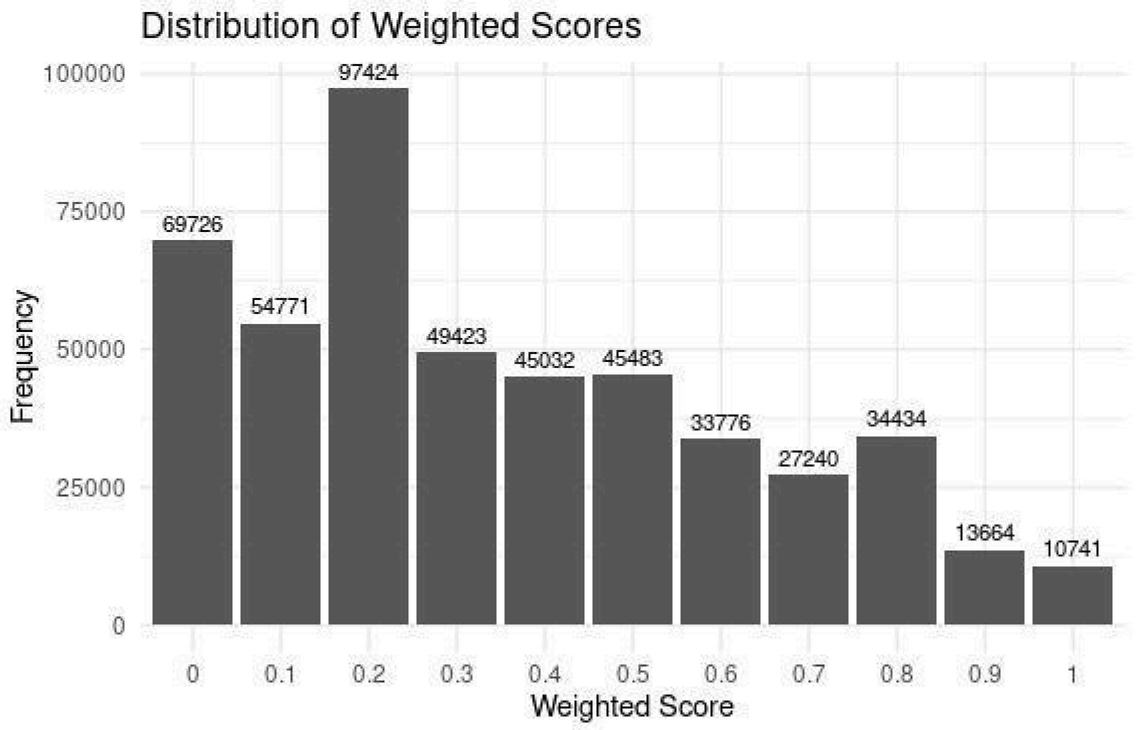


**Supplementary Figure 1: Barplot of the Distribution of Neuroticism Scores in UKB**

Neuroticism scores were calculated following Nagel et al. (2018)'s exclusion criteria:

- Individuals with more than 2 invalid responses were excluded.

For the remaining participants:

- Weighted scores = Sum of all responses/Total number of valid responses
- Scores ranged from 0 to 1.

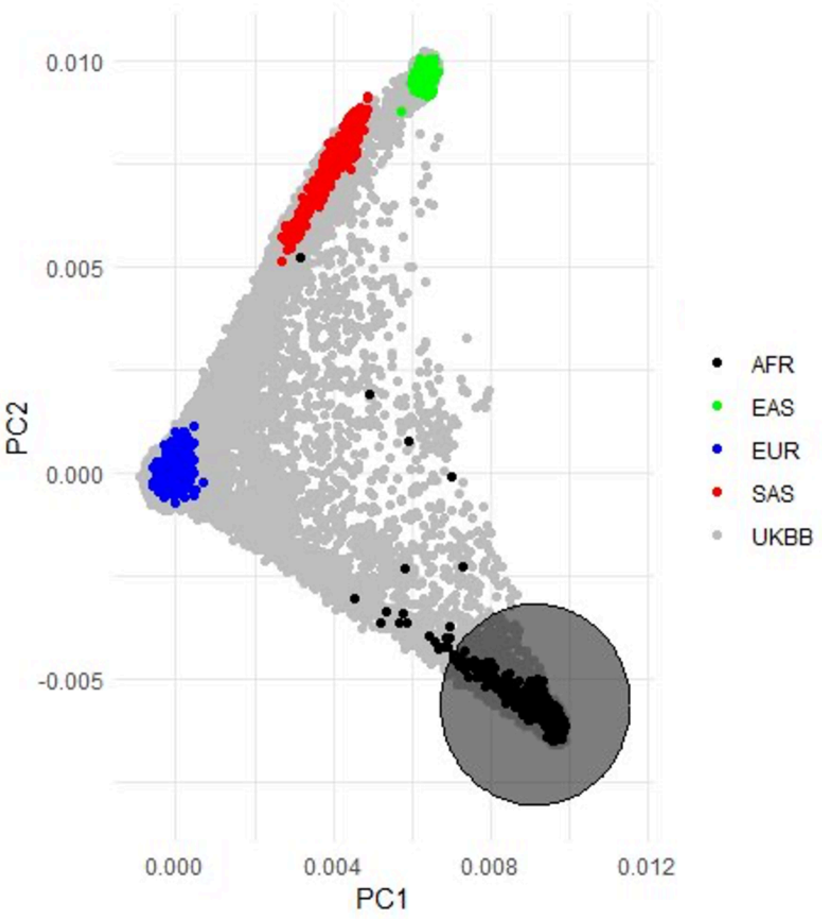


**Supplementary Figure 2: PCA plot of 1000 Genomes individuals projected on UKB samples.**

The PCA analysis was performed using TeraPCA. The circle radius shows the 3 SD from the mean of the 1kG AFR for the first 3 PCs, so all UKB individuals who fall in the circle were assigned as AFR in the PCA step. The same approach was repeated for the other ancestries.

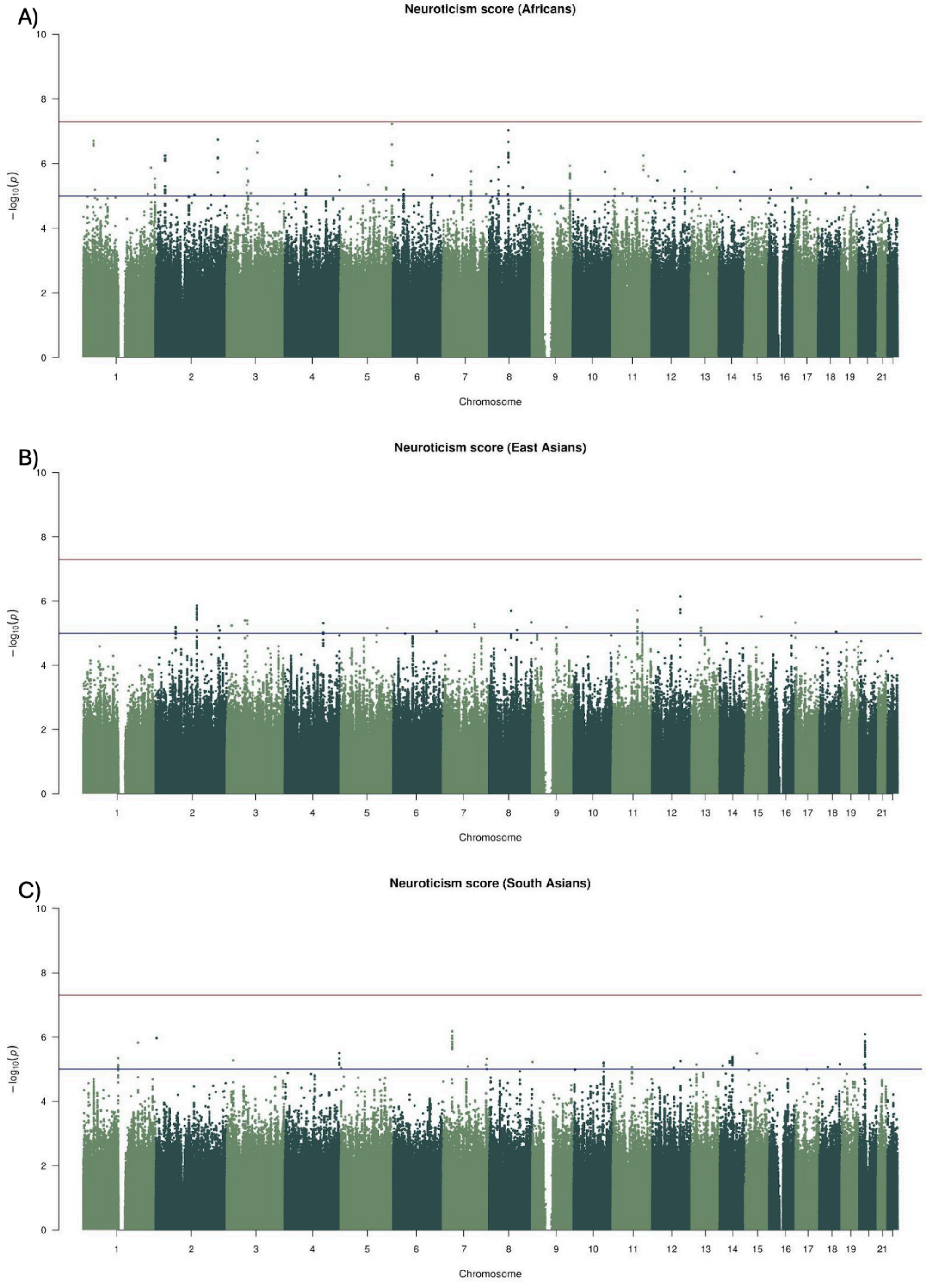


**Supplementary Figure 3: Manhattan Plots of Individual GWAS in UK Biobank individuals.**

A) Manhattan plot of 6374 UK Biobank individuals of African ancestry. B) Manhattan plot of 1600 UK Biobank individuals of East Asian ancestry. C) Manhattan plot of 6007 UK Biobank individuals of South Asian ancestry. Red line indicates the genome-wide significant threshold $p=5x10^{-8}$, blue line indicates the potential significant threshold $p=10^{-5}$.

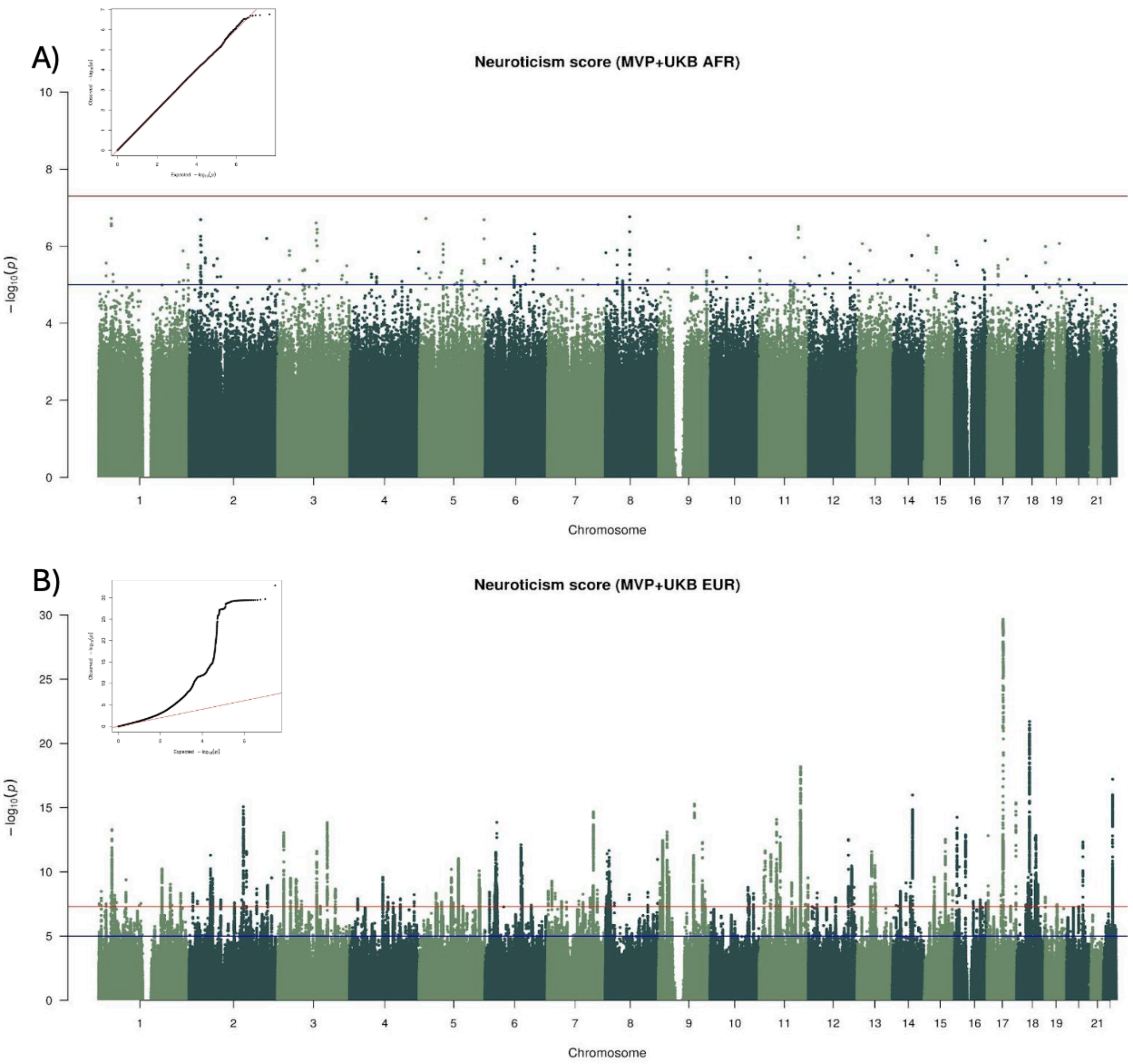


**Supplementary Figure 4: Manhattan and qq plots of Multi-ancestry analyses**

A) Meta-analysis of 37,691 MVP and UKB individuals of African ancestry. B) Meta-analysis of 623,482 MVP and UKB individuals of European ancestry. The meta-analyses were performed using METAL. Red line indicates the genome-wide significant threshold $p=5x10^{-8}$, blue line indicates the potential significant threshold $p=10^{-5}$.

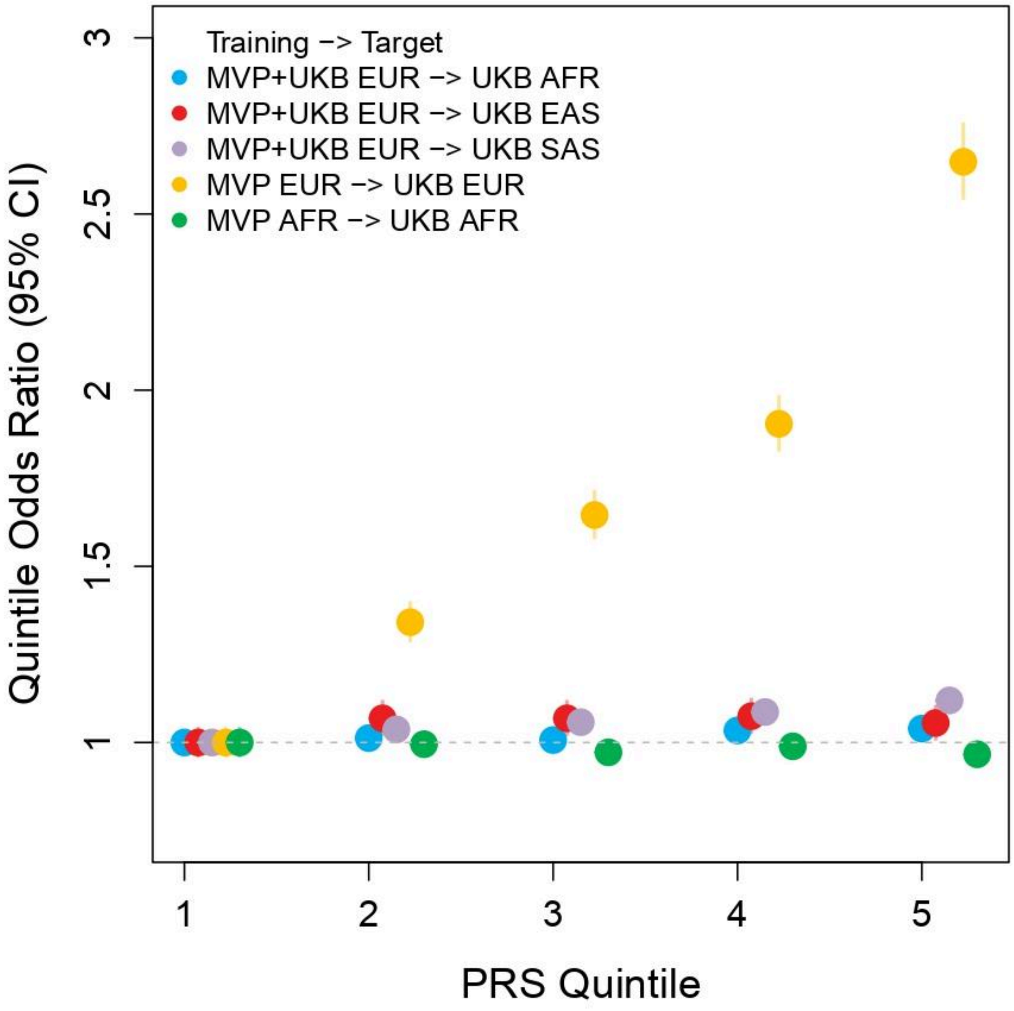


**Supplementary Figure 5: Within-ancestry and cross-ancestry PRS associations of Neuroticism.**

The y axis represents Neuroticism (NEU) risk relative to the lowest quintile of PRS with 95% CIs. For the **Within-ancestry**, predictions based on MVP EUR training data (yellow circles) demonstrate a significant performance increase in EUR samples compared to predictions based on the MVP AFR (green) counterpart. For the **Cross-ancestry**, Predictions based on MVP+UKB EUR training data perform poorly across all target datasets.

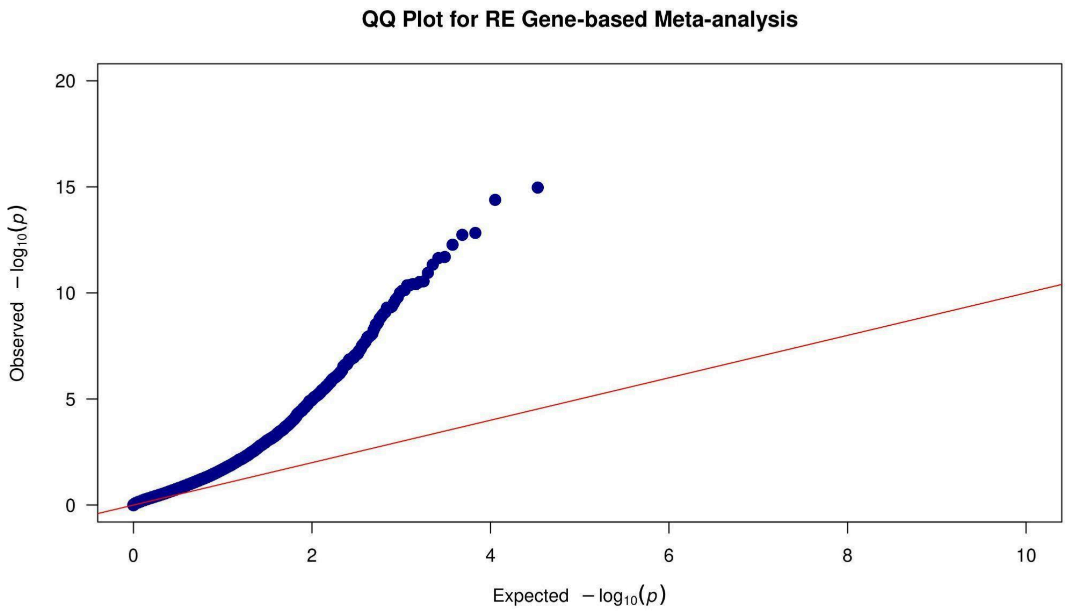


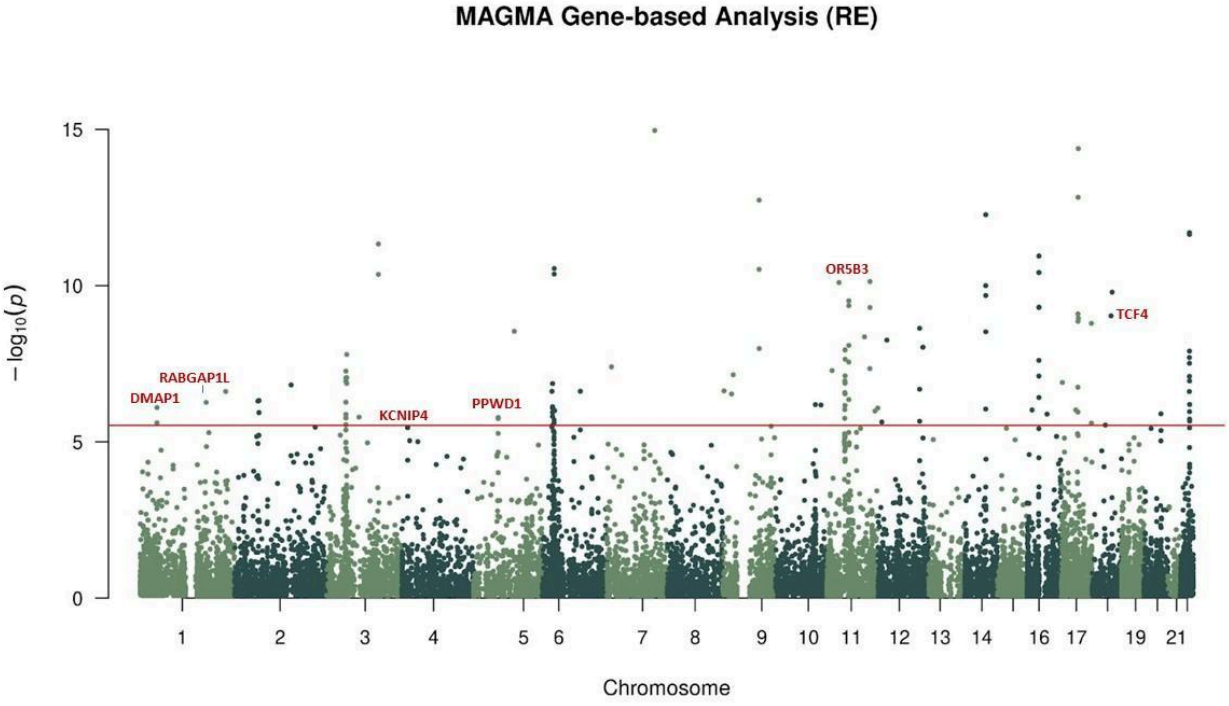


**Supplementary Figure 6: Manhattan and qq plots of gene-based analysis using MAGMA**

Manhattan and quantile plots of the MAGMA gene-based analysis based on summary statistics from the multi-ancestry GWAS. The results show a normal distribution curve with the novel genes highlighted in red.

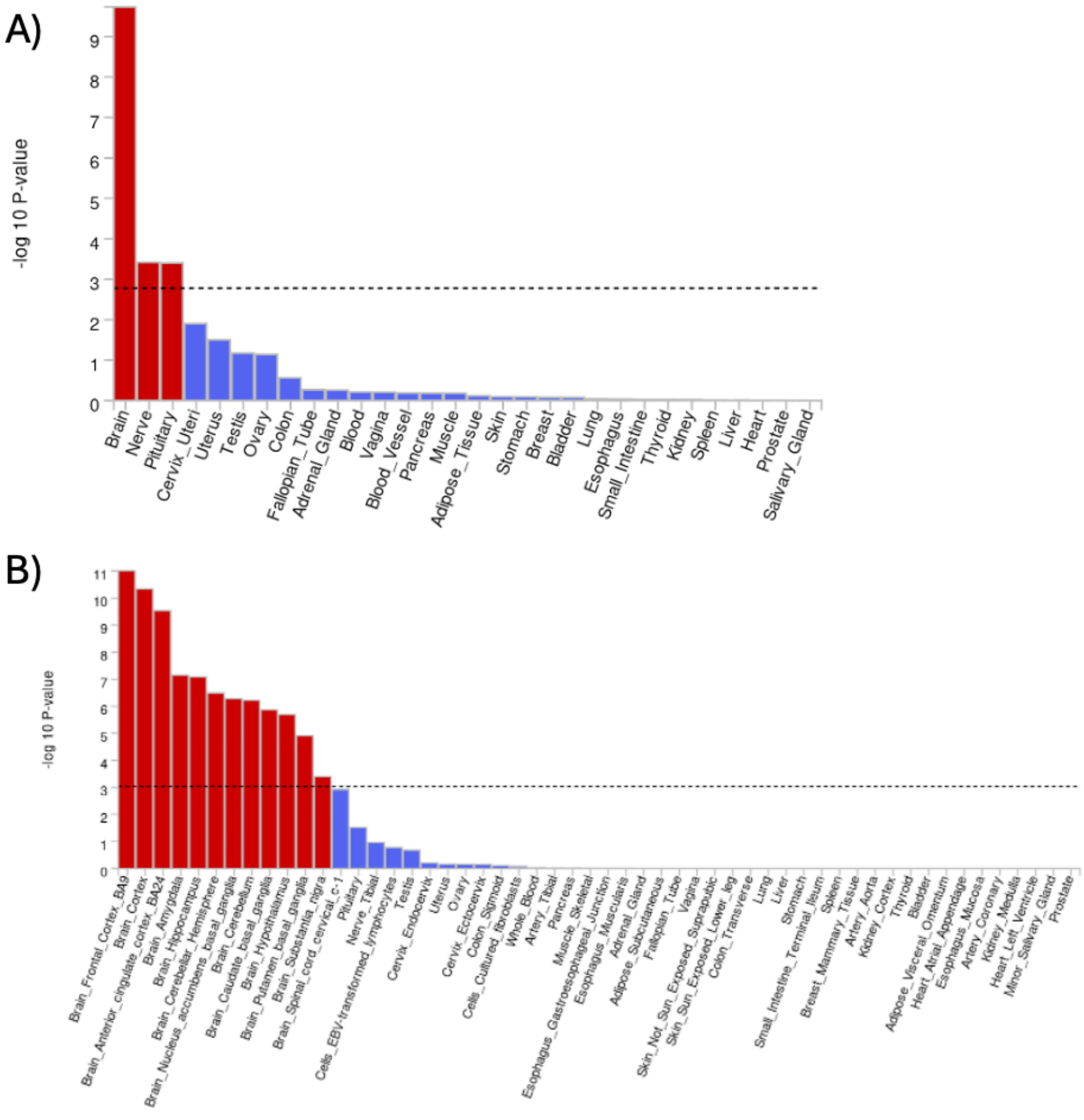


**Supplementary Figure 7: Tissue enrichment analysis in FUMA.**

[A].Bar charts show the enrichment of the brain, nerve, and pituitary regions in the gene expression analysis done using 30 broad tissues

[B]. Bar charts show the enrichment of brain tissues using 53 specific tissues from the online tool-FUMA.

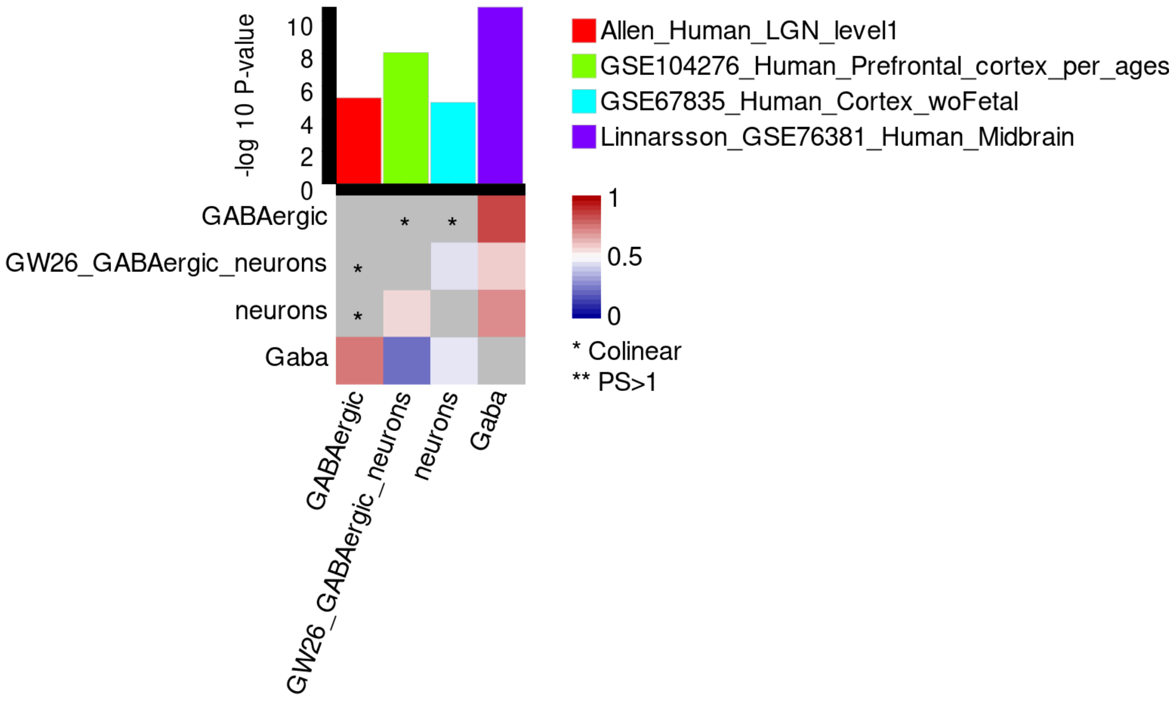


**Supplementary Figure 8: FUMA Cell-type results.**

Using different tissue cell types from FUMA, the heatmap showcases the cell-types that are enriched for Neuroticism after multiple correction testing is done.
